# Digital monitoring and action planning to reach zero-dose and under-immunised children: Leveraging data for targeted immunisation responses

**DOI:** 10.64898/2026.03.03.26346932

**Authors:** Mariam Zahid Malik, Naeem uddin Mian, Zahid Memon, Muhammad Wasim Mirza, Umer Farooq Rana, M. Adeel Alvi, Wardah Ahmed, Asma Ummad, Ammara Ali, Usama Naveed, Khayyam Saeed Malik, Muhammad Sufyan Chaudhary, Maria Waheed, Amna Sattar

## Abstract

**Background:** Persistent inequities in immunisation coverage, particularly among zero-dose and under-immunised children, continue to challenge Pakistan’s Expanded Programme on Immunisation. Weak feedback loop, inconsistent data quality, and limited real-time monitoring impede effective decision-making. This Implementation Research was conducted under the MAINSTREAM Initiative funded by Alliance for Health Policy and Systems Research (AHPSR) and supported by the Aga Khan Community Health Services Department and National Institutes of Health Pakistan to design, implement, and evaluate a digital monitoring and action planning tool to strengthen data-driven decision-making within routine immunisation systems.

**Methodology/Principal Findings:** A co-creation approach was employed to design a digital monitoring solution through inclusive consultations, key informant interviews, and focus group discussions with EPI Punjab at provincial and district levels. The solution included a customized mobile application for data collection and a Power BI visualization dashboard to map low-coverage areas, identify drivers of dropouts and zero-dose children, and capture caregivers’ information sources to inform targeted communication. The intervention was piloted in 60 households across six clusters of a Union Council of District Lahore. Advanced analytics identified reasons for non-vaccination and missed opportunities, generating tailored recommendations and practical plans for program managers. The analysis assessed acceptability, adoption, fidelity, and perceived scalability through field observations, system use, and stakeholder feedback.

The co-developed digital tool enhanced visibility of coverage gaps through UC-level mapping, real-time dashboards, and structured action planning. Pilot testing in Lahore showed strong acceptability, ease of use, fidelity, and adaptability among managers, supervisors, and vaccinators. Scalability and sustainability potential were demonstrated, though barriers included leadership turnover, system fragmentation, workload pressures, and resource constraints.

**Conclusion:** The tool demonstrated feasibility to strengthen immunisation equity, accountability, and responsiveness. Co-creation with stakeholders enhanced ownership, operational relevance, and adoption, while complementing existing platforms. Sustainability will depend on effective integration, local ownership, capacity building, and accountability, while scalability requires interoperability, resource commitment, policy support, and alignment with existing workflows.

**Author Summary:** Many children in Pakistan still miss routine vaccinations, especially those who have never received any vaccines or who drop out before completing the schedule. These children are often invisible within routine reporting systems, making it difficult for health managers to identify gaps and respond effectively. In this study, we developed and evaluated a digital monitoring and action planning tool designed to help immunisation managers and frontline workers better identify and respond to these gaps.

We worked collaboratively with provincial and district immunisation staff to co-design a user-friendly system that combines mobile data entry with interactive dashboards for supervision and planning. The tool was piloted in an urban district of Lahore, where health workers and managers reported high acceptability and ease of use. The system helped improve visibility of missed children, supported follow-up actions, and strengthened accountability across different levels of the immunisation program.

Our findings show that digitally enabled, co-created tools can strengthen routine immunisation systems by improving data use for action, supporting more responsive service delivery, and promoting equity. This work offers practical insights for scaling digital solutions to reach underserved populations and improve immunisation performance in similar low- and middle-income settings.

## 1. Introduction

Immunisation is globally recognized as one of the most cost-effective public health interventions, preventing millions of deaths each year [1]. Yet persistent inequities in vaccine coverage undermine progress in many low and middle-income countries, including Pakistan. Despite the launch of its Expanded Programme on Immunisation (EPI) in 1978 and the introduction of multiple new vaccines, challenges such as workforce shortages, cold chain gaps, data quality issues, and vaccine hesitancy continue to limit impact [2]. According to the Third-Party Verification Immunisation Survey (TPVICS) 2021, 76% of children aged 12–23 months are fully immunised, whereas an increase of around 1-2% (78%) was reported in 2023 [3].

Punjab, the country’s most populous province, has emerged as a leader in strengthening immunisation performance. It reported nearly 90% full immunisation coverage in 2023 [4] and pioneered key governance reforms, including integration of EPI financing into regular government budgets and stronger accountability through quarterly reviews. Punjab has also led digital health innovations such as e-Vaccs, which raised vaccinator attendance from ∼22% to 92% in 3 years and sharply improved coverage [5,6,7] alongside NEIR/EMR registry [8,9,10], EPI-MIS (Gavi Targeted Country Assistance Plan), Health Information and Service Delivery Unit (HISDU) GIS Portal and the vLMIS, which together enhanced monitoring, planning, and supply chain efficiency [11,12,13,14].

Nevertheless, disparities remain, findings from a 2024 – Rapid Cluster Coverage Assessment (RCCA) in Lahore and Faisalabad revealed overall Pentavalent-3 coverage of 91% but significant district gaps, with full immunisation reaching 93% in Lahore versus only 57% in Faisalabad. Zero-dose prevalence stood at 3%, while misconceptions, inconvenient vaccination timings, and low maternal tetanus-diphtheria coverage (10%) highlighted persistent demand-side barriers.

To address these systemic gaps, the Aga Khan University (AKU), under the MAINSTREAM (Institutionalizing learning by mainstreaming embedded implementation research in country immunisation programs) Initiative supported by the Alliance for Health Policy and Systems Research (AHPSR), WHO, and Gavi, partnered with Contech International to design and pilot a Digital Monitoring and Action Planning Tool. This tool was conceived to provide real-time data, enable geo-spatial mapping of zero-dose clusters, and support targeted action planning. Developed through embedded implementation research, it emphasized co-creation with stakeholders to ensure alignment with local needs and readiness for long-term adoption and scalability.

### 1.1 Research Workflow and Implementation Framework

The implementation process was carefully structured into iterative phases (Fig 1). It began with the formation of a multi-disciplinary team comprising public health experts, IT specialists, immunisation managers, and field supervisors. To ensure methodological rigor, AKU convened a workshop from 8^th^-9^th^ October 2024 in Karachi to refine research protocols and strengthen the study design. This was followed by an inception meeting with provincial and district EPI teams to clarify objectives, build consensus, and finalize protocols. Together, the following steps aligned the expectations of researchers and implementers while laying a strong foundation for methodological rigor and collaborative ownership (Fig 1**: Steps followed for digital tool implementation**).

**Fig 1:**
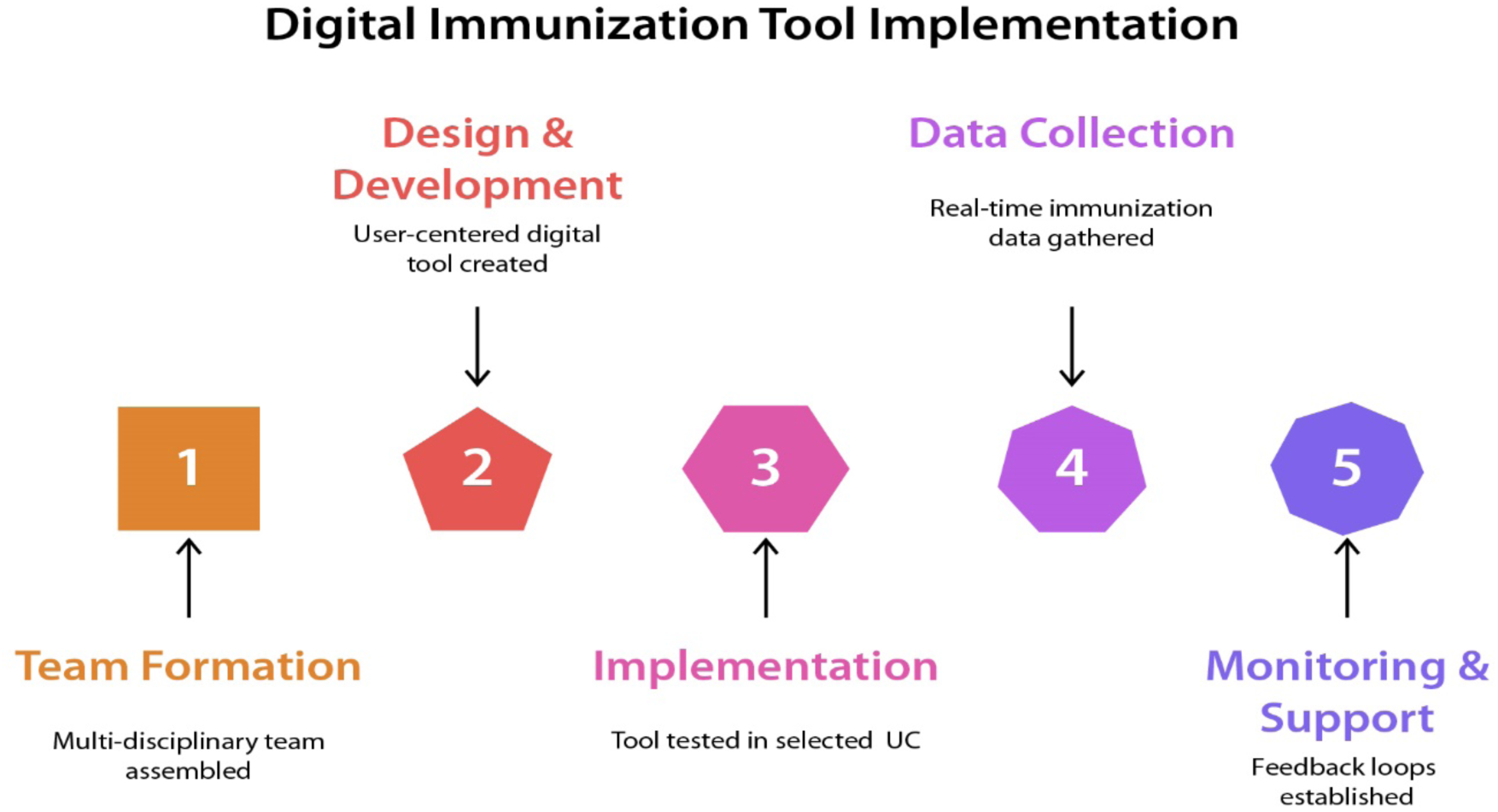
Steps followed for digital tool implementation.

#### 1.1.1 Development phase

The research team prioritised desk review followed by co-creation of user-centred design for the digital tool. The digital monitoring tool was developed by Contech’s Digital Team. The tool development consisted of two core components: (1) Development of a mobile application designed for vaccinators to capture household and child-level data in real time; and (2) Visualisation of data on a Power BI–based dashboard for supervisors and managers to depict coverage gaps, generate action plans, and track progress. The design was refined iteratively through stakeholder feedback and real-time piloting. The desk review underscored Punjab’s success in leveraging digital innovation and accountability mechanisms to strengthen EPI, while also highlighting enduring challenges in equity, awareness, and sustainability. Strategic scale-up of digital solutions, stronger community engagement, and consistent periodic assessments like RCCA were considered essential for closing immunisation gaps. Stakeholders were actively engaged through structured consultations, demonstrations of early prototypes, and iterative feedback sessions. This process revealed the practical requirements of vaccinators, such as the need for offline functionality, GPS tagging, and simplified data entry, alongside managerial requirements like geo-spatial mapping and automated action planning. These insights were translated into technical requirements, guiding the development of the mobile application and Power BI dashboard.

#### 1.1.2 Implementation phase

During this phase, the digital tool was piloted in a Union Council of Lahore District, following WHO criteria for a Rapid Coverage Cluster Assessment (RCCA). Employing Lot Quality Assurance Sampling (LQAS) [15] technique, 06 clusters were identified within the UC using Probability Proportional to Population Size (PPS), and from each cluster, ten households were selected through yielding a total of 60 households. Eligibility required the presence of a child under two years of age and a parent or caregiver. Enumerators received orientation and hands-on training in the use of the mobile application, while district managers were guided on interpreting and applying dashboard analytics. These capacity building sessions combined didactic presentations with practical exercises, ensuring that both frontline workers and managers gained confidence in using the tool. Using the customised mobile android app, real-time digital data was collected on routine immunisation status of children aged 0–23 months and tetanus-diphtheria (TD) vaccination of mothers. Immediate visualisation of zero-dose and under-vaccinated children, along with mapping of coverage gaps, provided actionable insights.

The tool’s action-planning feature further enabled supervisors to assign tasks, set deadlines, and monitor follow-up actions, fostering accountability and timely corrective measures. Monitoring and fidelity tracking were critical to assessing implementation integrity. Field visits were carried out to observe data collection processes, identify operational challenges, and provide real-time troubleshooting support. Structured feedback loops ensured that issues such as syncing delays, limited digital literacy, and data entry errors were addressed promptly. This phase highlighted the adaptive capacity of the intervention, with refinements such as the introduction of contextual tooltips, improved UC selection logic, and interface adjustments for mobile devices implemented rapidly in response to user feedback.

#### 1.1.3 Dissemination phase

This phase marked the conclusion of the pilot cycle. A formal dissemination workshop was organised under the Mainstream Initiative in the federal capital, jointly hosted by the National Institute of Health (NIH) and the Federal Directorate of Immunisation (FDI) in collaboration with Aga Khan University (AKU) under the funding of AHPSR. The event was attended by EPI focal persons from across the provinces, ensuring national-level representation. The workshop provided a platform not only to share key findings but also to secure policy-level endorsement for potential scale-up. Stakeholders actively contributed feedback on sustainability pathways, integration with existing platforms such as NEIR and DHIS2, and alignment with provincial immunisation strategies. By fostering dialogue and consensus, this phase consolidated stakeholder ownership and laid a strong foundation for scaling the tool beyond the pilot setting.

## 2. Results

The implementation of the digital monitoring and action planning tool yielded multiple outcomes. The most notable achievement was the significant improvement in the visibility of coverage gaps. Through the mobile application, vaccinators were able to capture household-level immunisation data in real time, which synchronised seamlessly with the Power BI dashboard. This allowed managers to monitor coverage trends at the Union Council level, identify zero-dose and under vaccinated clusters, and initiate timely interventions. Stakeholders consistently underscored the tool’s acceptability and adaptability. Provincial EPI officials emphasised its utility in providing real-time insights for decision-making, while district-level managers valued its ability to pinpoint problem areas with precision. Vaccinators, who had traditionally relied on paper-based systems, reported that the app simplified their workflow, reduced paperwork, and enhanced accuracy. A vaccinator described the application as *’a tool that makes work easier and more transparent.’*

Fidelity tracking demonstrated that the intervention was largely delivered as intended. Field observations indicated that vaccinators were consistently able to record data accurately, and that supervisors were actively using the action planning module to assign follow-up tasks. The integration of accountability mechanisms into the dashboard enabled supervisors to track task completion rates and provide targeted support to vaccinators in underperforming areas. This was perceived as a major step forward in strengthening accountability within the EPI system.

The pilot also revealed several challenges. Initial resistance to adopting digital tools was observed among some staff, particularly those with limited exposure to technology. Leadership transitions during the initial period of project caused delays in decision-making and reduced continuity. Nevertheless, the overall sentiment among stakeholders was positive, with many highlighting the tool’s strong potential for scale-up if enabling environment is provided.

### 2.1 Results – Development & Deployment of Digital Monitoring and Action Planning Tool

As part of implementation research to strengthen immunisation coverage and evidence-based decision-making, a mobile-responsive Digital Monitoring & Action Planning Tool was piloted in a district setting. Designed to be intuitive, scalable, and compatible with existing health systems, the tool integrates a mobile application for field staff with a dashboard for district and provincial managers. The tool enables role-based access, interactive visualisations, GIS mapping of zero-dose clusters, and analytics to identify reasons for missed vaccinations, while its action-planning module supports task assignments, timelines, and progress tracking. Collectively, these features enhance transparency, drive timely action, and ensure effective monitoring at all administrative levels.

#### 2.1.1 Tool Design and Development Process

The digital monitoring tool has been developed using *Microsoft Power BI* to enhance the effectiveness and transparency of field data collection and monitoring processes for immunisation programs. The backend is powered by a *MySQL database*, ensuring structured, reliable, and scalable data storage. Power BI connects directly to this database to retrieve and visualise required data. Once developed, the tool was published on the Power BI Service, enabling secure access through both web and mobile platforms.

The design phase began with a thorough requirements-gathering exercise engaging field teams, program managers, and data analysts. Key features were mapped based on user needs, which included offline data entry, location tagging, and visual performance tracking. Development was executed in iterative phases:

- **Phase 1**: Creation of MySQL database schemas and API endpoints
- **Phase 2**: Web dashboard in Power BI using Import Mode data connection.
- **Phase 3**: Publishing the dashboard on the power bi services.

#### 2.1.2 Dashboard and Mobile App – Key Features

##### 2.1.2.1 Data Filtration

Users can dynamically filter data by province, district, UC, and child’s gender to customise insights and prioritise analysis (Fig 2).

**Fig 2:**
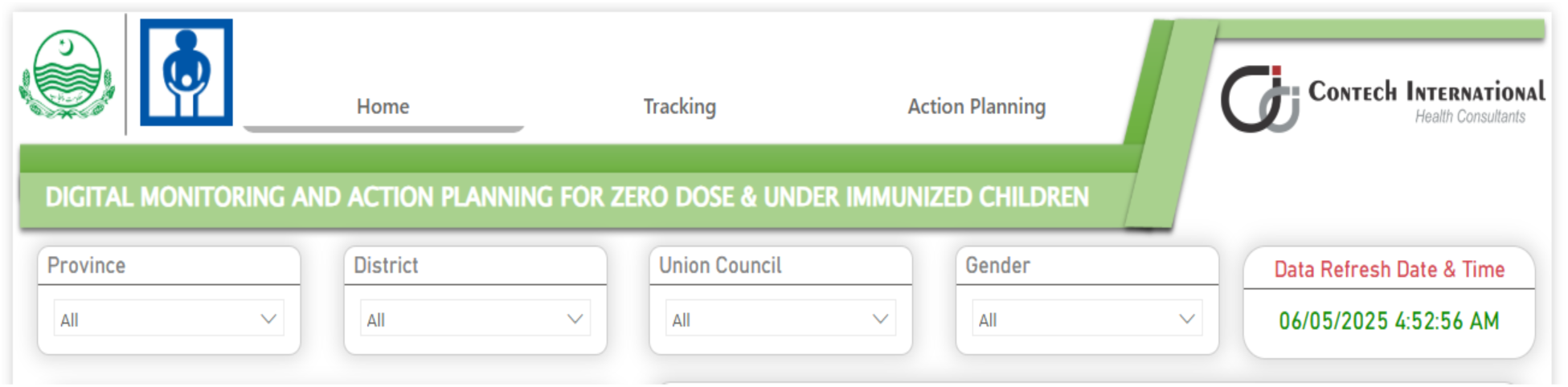
Dashboard interface for data filtering and last update time.

##### 2.1.2.2 Basic Information for child and the mother

Key information about children categorised as zero-dose, never vaccinated, or currently due for immunisation are reflected in the dashboard. It is followed by detailed data on the child’s mother and the corresponding union council (UC) information (Fig 3).

**Fig 3:**
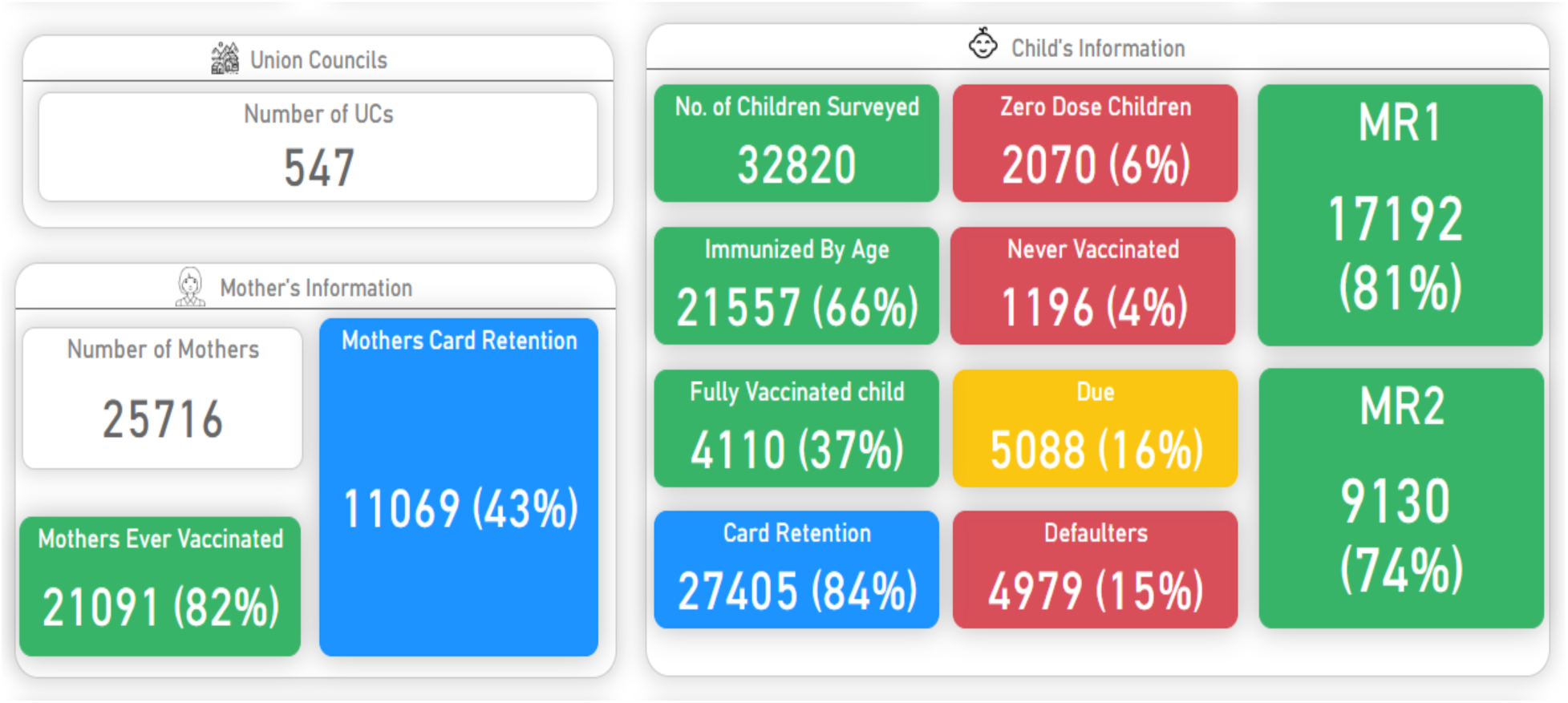
KPI tiles for UC coverage and maternal/child immunisation performance.

##### 2.1.2.3 Map

An interactive map displays information based on the user’s selection of a province or by hovering over highlighted areas (

Fig **4****: *Province map with* immunisation coverage and child card retention details**). The tooltip provides key details including the province name, the number of UCs within the province, the count of zero-dose children and the number of never vaccinated children.

**Fig 4:**
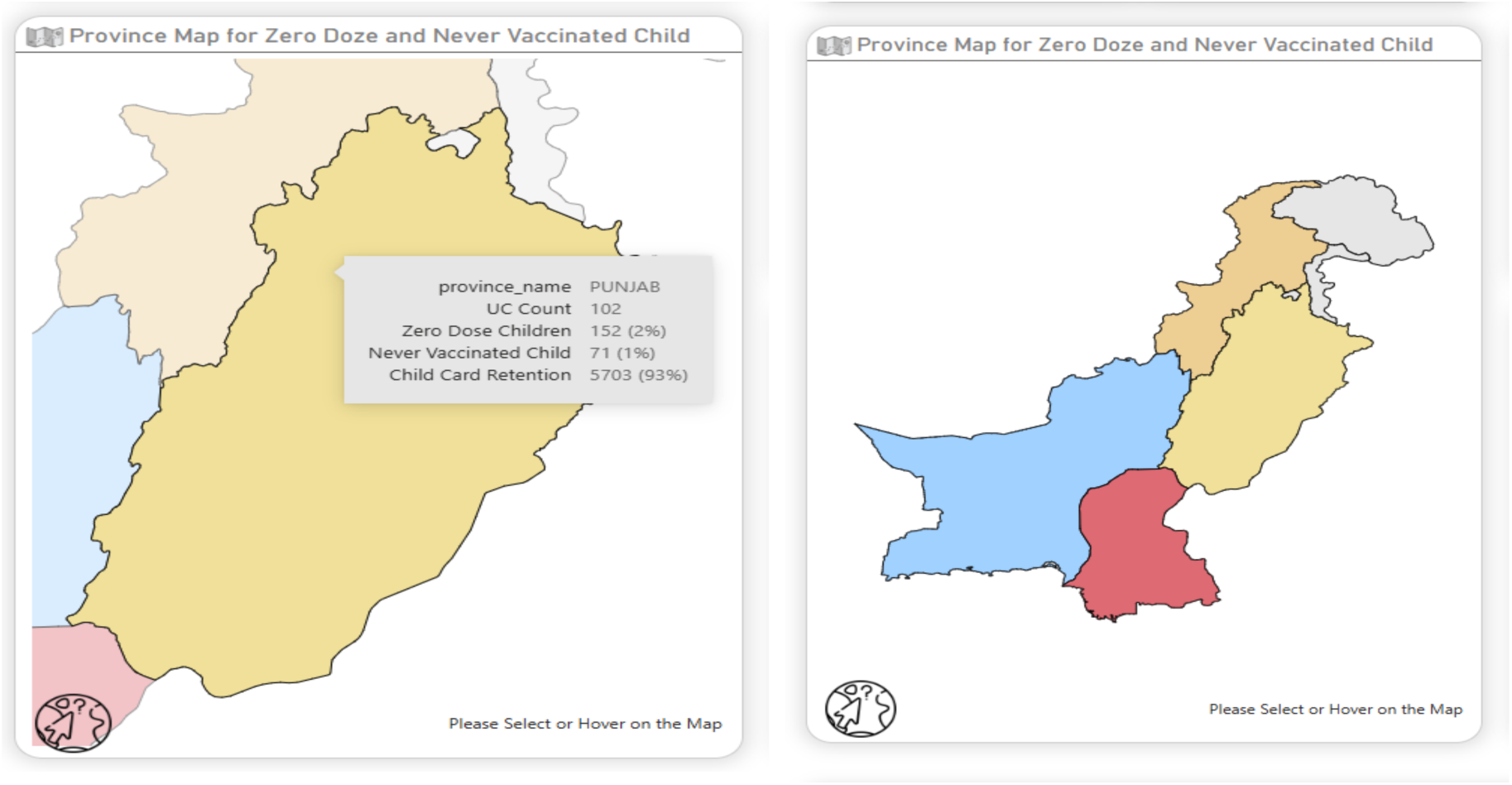
**Province *map with* immunisation coverage and child card retention details**

##### 2.1.2.4 Action Planning

The Action Planning within the dashboard is designed to guide targeted interventions based on the number of zero-dose children identified in each union council. This component enables EPI teams and decision-makers to prioritise areas requiring attention and tailor their response accordingly. Utilising the LQAS technique, following thresholds have been established to facilitate evidence-based decision-making:

#### 2.1.3 Threshold-Based Decision Support for Action Planning

The system categorises sample of 60 children/HH within one union council based on the number of zero-dose children and recommends specific actions:

○ 0–3 Zero-Dose Children (out of 60): When the count falls within this range, it is considered a low-risk area. No immediate action is required, but continued monitoring is advised to ensure numbers do not increase.
○ 4–8 Zero-Dose Children (out of 60): This range triggers the system to analyse and identify the most common reasons for non-vaccination in the area (e.g., child was sick, parental unawareness, access issues). Based on this analysis, the algorithm provides tailored suggestions to address these root causes, such as community awareness sessions, follow-up visits, or coordination with local health workers.
○ 9 or More Zero-Dose Children (out of 60): When the number exceeds this threshold, it indicates a high-priority area. The algorithm recommends conducting/redoing of a targeted vaccination campaign within the respective union council. This may involve deploying outreach teams, setting up mobile vaccination camps, or coordinating with community leaders to improve coverage. This tiered approach ensures efficient use of resources, focusing interventions where they are most needed and promoting proactive measures in emerging risk areas. It also helps in tracking the effectiveness of field strategies over time and improving overall immunisation coverage (Fig 5).

**Fig 5:**
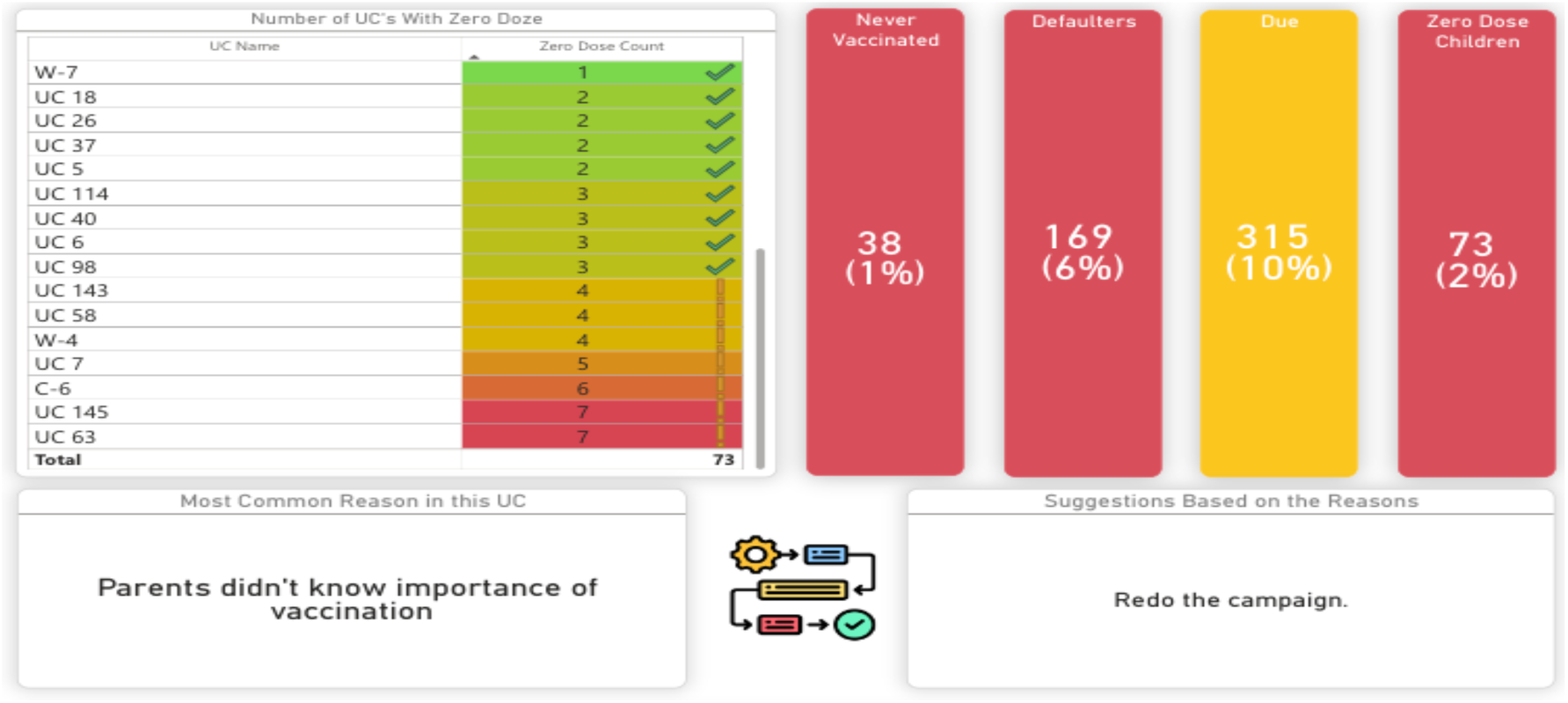
**Dashboard view of zero-dose UCs with reasons and suggested corrective actions**

##### 2.1.3.1 Information tree

This part of the dashboard presents the data in the form of a hierarchical tree, allowing users to drill down and explore the underlying reasons why a child was not vaccinated (Fig **6** and Fig ***7***).

**Fig 6:**
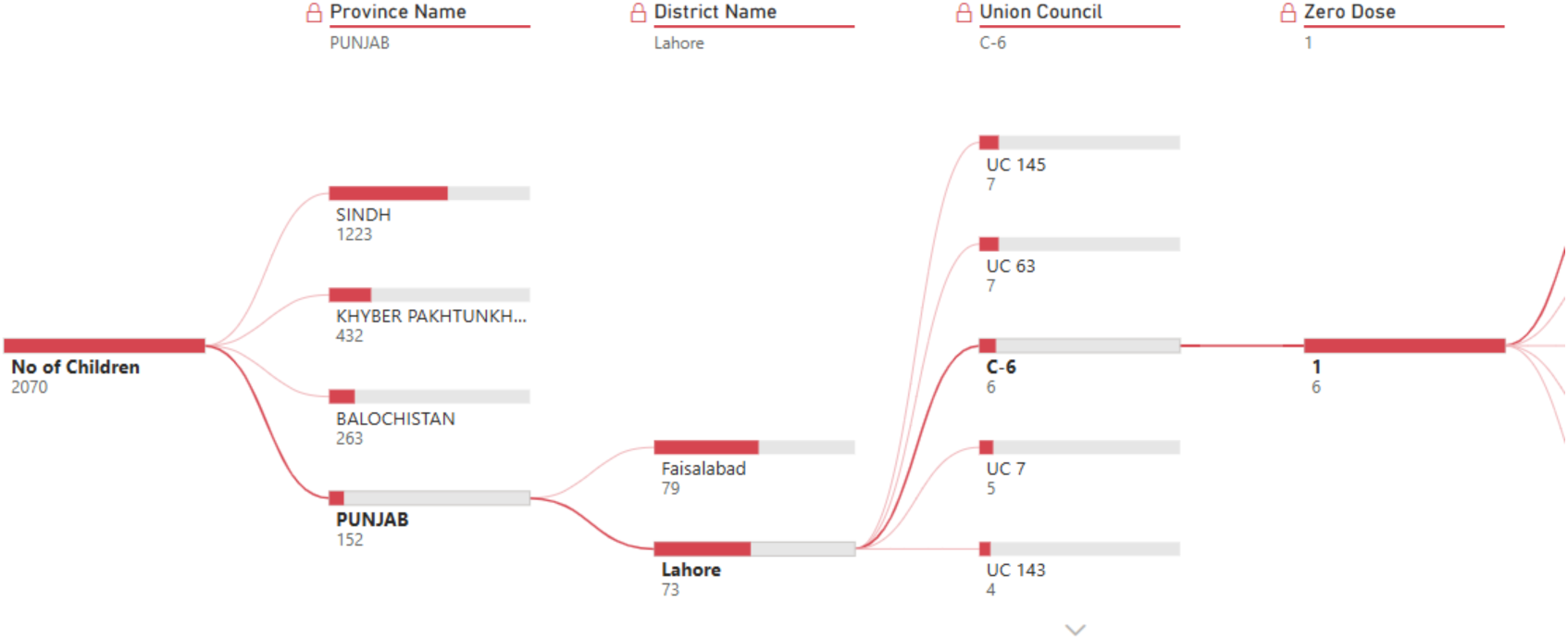
**Hierarchical tree visualisation for drilling down zero-dose children by province, district, and UC.**

**Fig 7:**
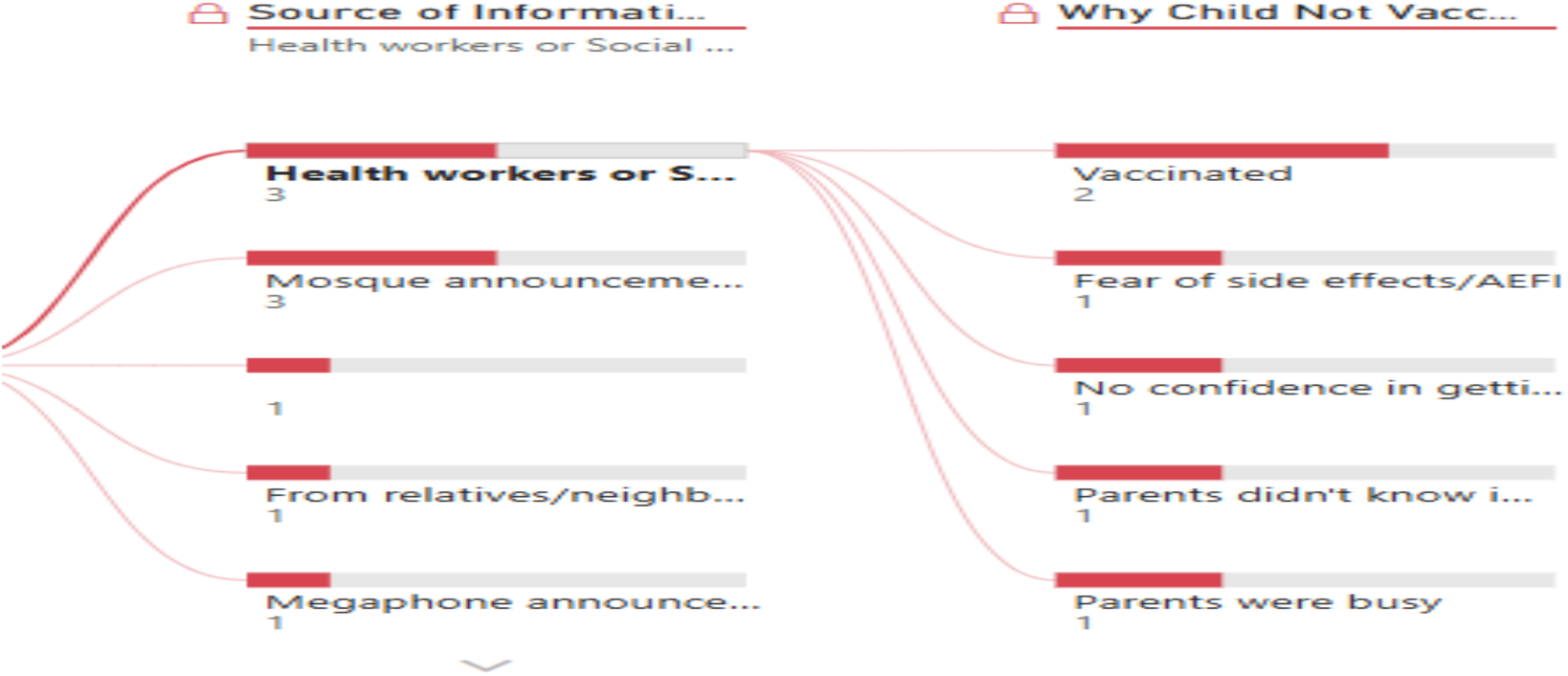
**Information tree linking sources of awareness to reasons for non-vaccination.**

##### 2.1.3.2 Map for child information

The dashboard includes an interactive mapping feature that visualises the geographic distribution of zero-dose children. It provides detailed information at multiple administrative levels – union council, district, province, and national, allowing users to identify specific locations, quantify cases, and compare patterns across regions. The map also enables drill-down exploration to highlight areas with higher burden and child identification and specific location supports data-driven targeting of immunisation efforts (Fig 8 Fig 9).

**Fig 8:**
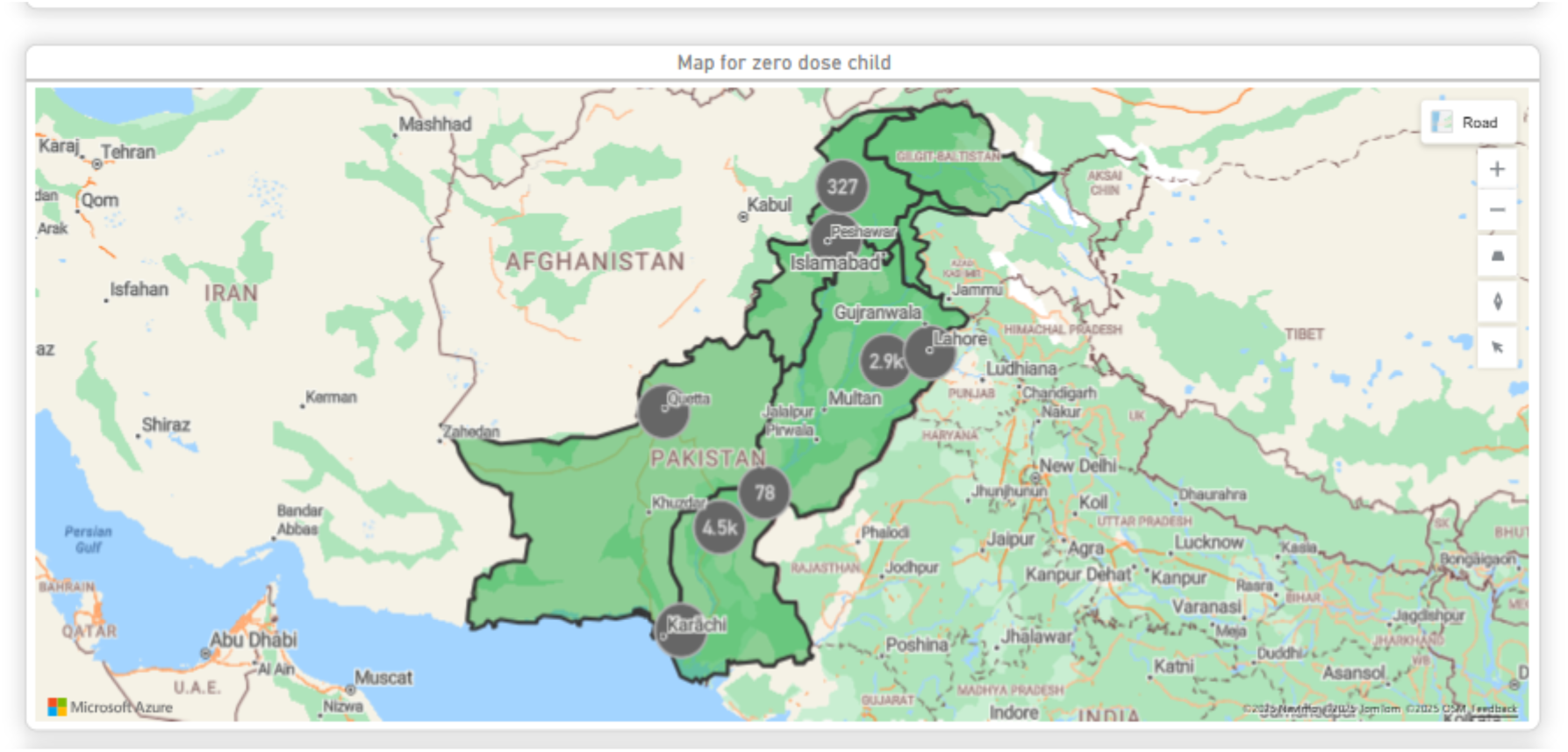
**Map showing province- and district-level distribution of zero-dose children**

**Fig 9:**
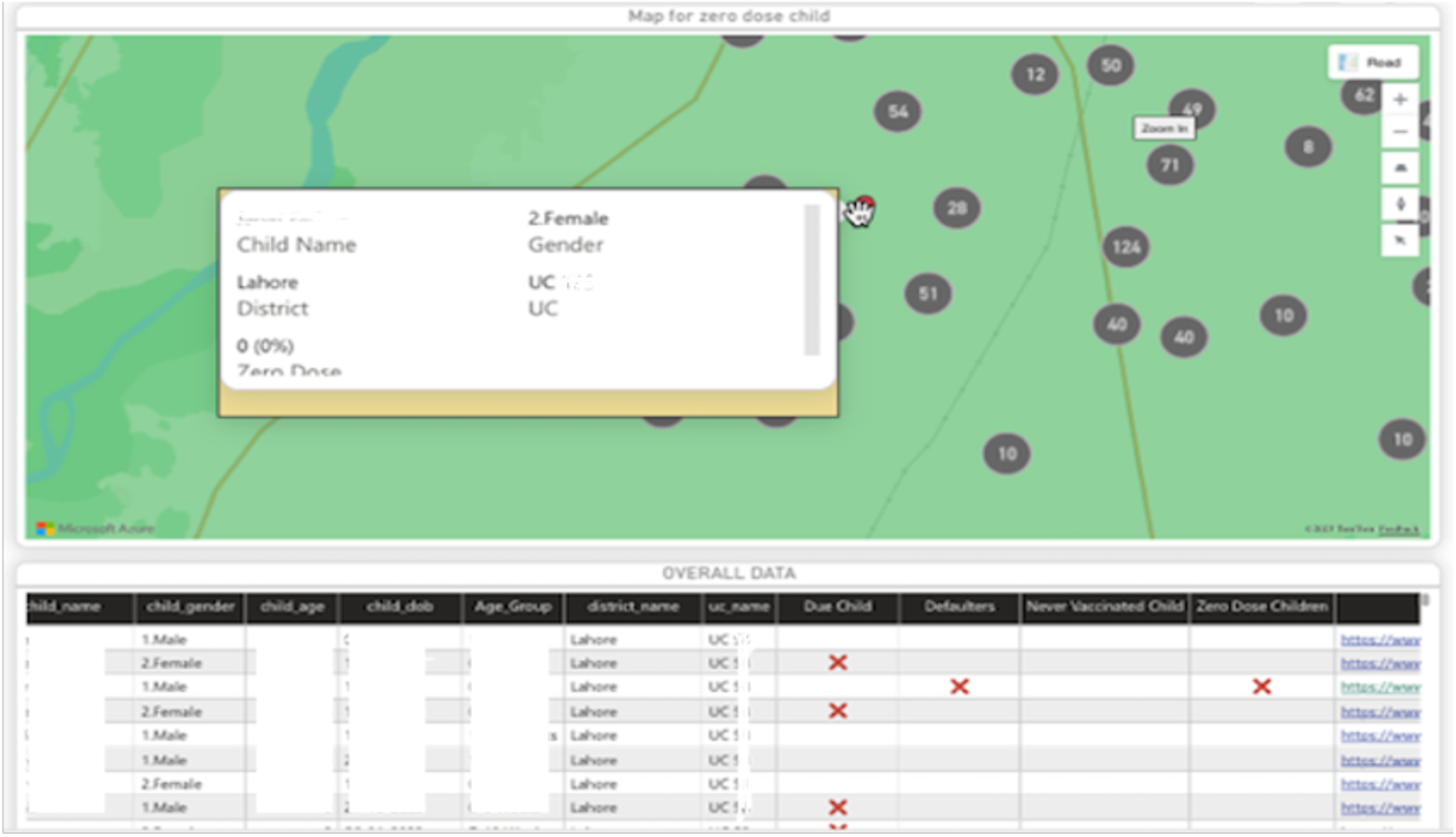
**Child identification and location at UC level**

##### 2.1.3.3 Tracking

This section of the dashboard is dedicated to tracking the actions suggested and taken at the UC level. It ensures transparency and accountability by allowing users to monitor the progress of recommendations provided for addressing zero-dose cases or other health service gaps.

- **UC-Based Action Display**: Actions and suggestions are displayed in a structured format, categorised by each UC. This allows users to easily view which areas have pending actions, what suggestions have been provided, and what steps have been taken.
- **User Control and Customisation**: To promote flexibility, users can edit the suggestions associated with a UC by clicking on the “Edit” button. This feature enables field staff or administrators to update action plans based on local context, new findings, or progress made on previous recommendations.
- **Real-Time Progress Monitoring**: The real-time progress monitoring feature enables dynamic planning and timely interventions by organising actions at the union council level. Users can update progress instantly, edit suggestions, and mark completion of planned activities through an interactive checklist. This functionality not only provides guidance but also empowers users to actively manage and adapt the implementation process. Additionally, a print option allows easy sharing of progress reports with field teams, ensuring transparency and accountability (Fig 10**: Real-time progress monitoring with UC-level action tracking**).

**Fig 10:**
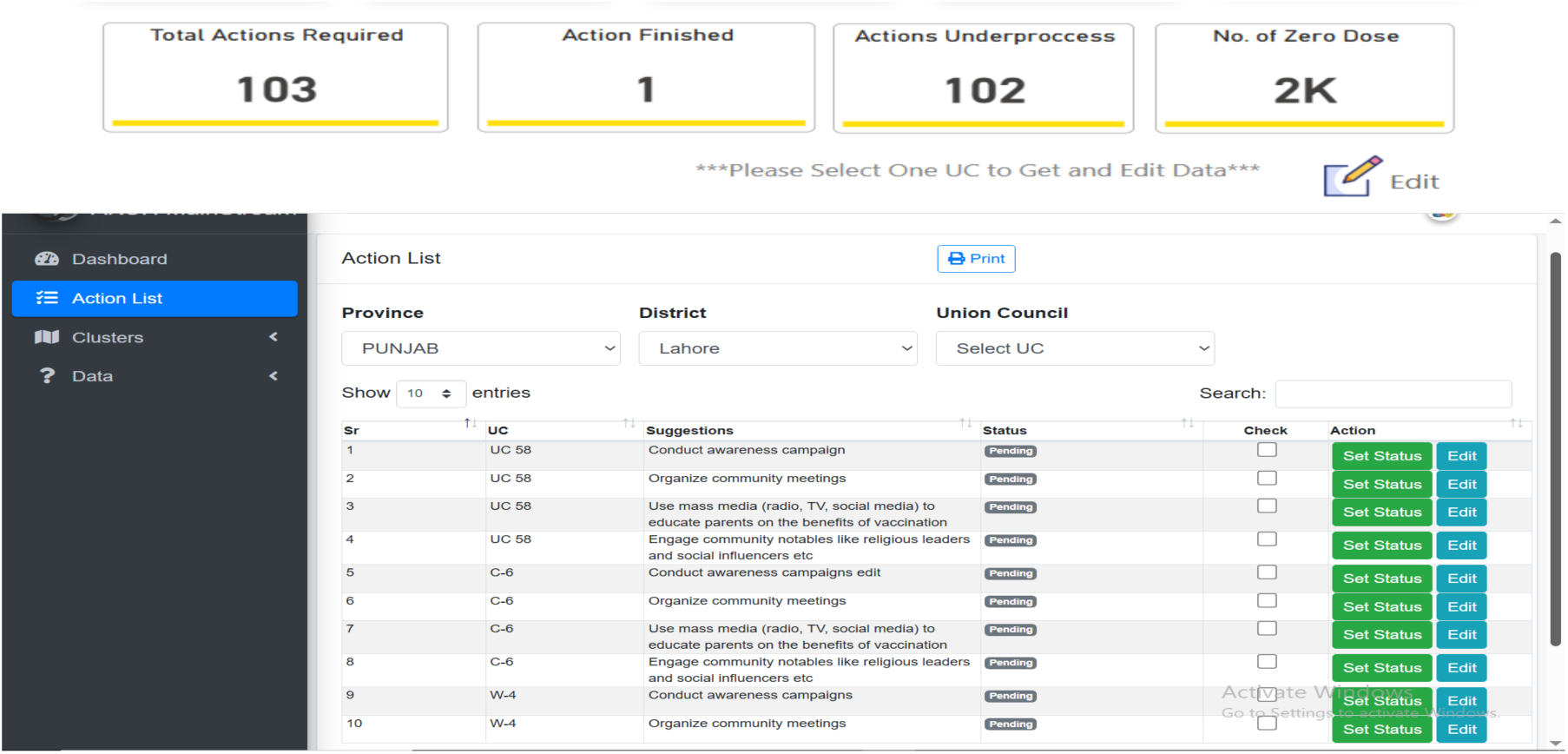
**Real-time progress monitoring with UC-level action tracking**

#### 2.1.4 Key Refinements

Stakeholder feedback from EPI managers, field teams, and technical staff guided several key refinements to improve usability and operational relevance. Improvements included optimising UC selection logic, adding tooltips for better understanding of key visuals, and simplifying mobile data entry to reduce errors and enhance field efficiency and printable checklist added in dashboard for dynamic planning and progress monitoring. These refinements directly addressed challenges encountered during early use. Originally built for desktop analysis, the Power-Bi dashboard was reconfigured for mobile responsiveness following feedback on the need for field accessibility. Enhancements included better navigation for small screens, real-time data syncing from the mobile app, and a 30-minute auto-refresh feature to maintain data accuracy. Visual demonstrations of the mobile interface supported wider adoption, reflecting a practical and adaptive design process. The refinements strengthened data visibility, accountability, timely decision-making and aid progress tracking of proposed actions, reinforcing the value of stakeholder-driven approaches in digital health interventions.

### 2.2 Results – Adoption, Adaptation, and Integration

#### 2.2.1 Provincial-Level Perspectives

Interviews with the Provincial Program Manager, EPI MIS Focal Person, and DHIS2 Coordinator revealed significant progress in Punjab’s immunisation coverage but highlighted persistent equity gaps. The Provincial Program Manager noted that “*despite nearing 90% coverage, disparities persist in urban slums, nomadic groups, and remote areas*.” The MIS Focal Person added that “*zero-dose children are concentrated in peri-urban and migratory belts; mobility and low awareness are major issues*,” while the DHIS2 Coordinator emphasised that “*inconsistencies in denominators remain a concern*.” All respondents recommended transforming Punjab’s digital immunisation platforms into a unified, real-time intelligence system through unique child identifiers linked to CNIC and birth registration, integrated GIS-enabled micro-planning, improved device connectivity, and strengthened data literacy.

##### 2.2.1.1 Micro-planning and Campaign Design

All informants reported that while micro-planning is institutionalised, it is hampered by outdated population data, migration trends, and offline functionality. The Provincial Program Manager highlighted that “*micro-plans need real-time population updates and seasonal migration insights*.” The MIS Focal Person stated that “*our micro-planning tools are strong but face underreporting and outdated denominators*,” and the DHIS2 Coordinator added that “*we use DHIS2 modules to monitor daily team performance and outreach, but offline syncing remains a challenge.*”

##### 2.2.1.2 Use of Data for Decision-Making

Data is routinely discussed in monthly review meetings, yet harmonisation and frontline data literacy remain weak. According to the Provincial Program Manager, “*monthly review meetings guide UC prioritisation, but data harmonisation is still weak.”* The MIS Focal Person noted that “*feedback loops are limited, and digital literacy is low at field level*,” while the DHIS2 Coordinator observed that “*we generate performance dashboards, but frontline use is hindered by low analytical capacity.*”

##### 2.2.1.3 Digitisation and Monitoring Tools

Punjab has deployed DHIS2, EMR, and E-Vaccs platforms; however, uptake remains inconsistent, particularly at peripheral levels. The Provincial Program Manager reflected that “*E-Vaccs and EMR have helped, but field-level uptake is inconsistent due to infrastructure limitations.*” The MIS Focal Person emphasised the need for “*tools that work offline, include GPS, and generate defaulter alerts automatically,”* while the DHIS2 Coordinator stated that “*DHIS2 needs unique child identifiers and better integration with NADRA and polio platforms.”*

##### 2.2.1.4 Capacity Building and Integration Needs

Informants emphasised the importance of workforce capacity and system interoperability. The Provincial Program Manager noted that “*training on data use and digital tools is essential for scale-up and sustainability.*” The MIS Focal Person added that “*district MIS cells need upgrades in both hardware and skills*,” while the DHIS2 Coordinator concluded that “*to evolve DHIS2 into an immunisation intelligence platform, we must invest in user trust and frontline ownership.”*

#### 2.2.2 District-Level Perspective

The District EPI Manager, Lahore, confirmed similar trends, focusing on operational challenges in addressing zero-dose and under-vaccinated communities. “*Our Lady Health Workers and outreach vaccinators play a critical role in flagging missed children, especially in peri-urban areas and urban slums*,” the Manager stated, but acknowledged that “*key barriers include vaccine hesitancy, urban migration, and inadequate human resources in informal settlements.”* The District Manager advocated expanding mobile teams, integrating digital newborn tracking with community surveillance, and partnering with telecom and technology providers. “*We need capacity building of vaccinators and data staff, reliable internet access, and integration of tools into existing systems*,” the respondent said. “*Digital monitoring can help by providing real-time feedback and reducing manual errors.”*

##### 2.2.2.1 Micro-planning and Campaign Design

The District EPI Manager reported that microplans are developed and validated at both UC and district levels, targeting katchi abadis and nomadic populations. “*Maps are marked with missed clusters based on last campaign data to improve planning,*” the respondent explained. However, “*common challenges include cold chain maintenance during mobile outreach, staff shortages, and insufficient time for social mobilisation.*”

##### 2.2.2.2 Evidence-Based Decision-Making

Data triangulation remains limited. “We analyse immunisation data at monthly review meetings to assess UC-wise performance,” the respondent stated, adding that “paper-based reporting adds to workload and reduces timeliness.”

##### 2.2.2.3 Digitisation and Monitoring Tools

The district uses EMR, E-Vaccs, and tablets mainly for attendance and session tracking. “*We would prioritise real-time data capture, GIS mapping, and dashboards for supervisors*,” the Manager noted, emphasising that “*training, IT support, and device maintenance are necessary for sustainable rollout*.”

Both provincial and district perspectives align on major challenges, including vaccine hesitancy, data fragmentation, outdated micro-plans, and weak frontline analytical capacity. Provincial informants emphasised system-level integration, interoperability, and predictive analytics, whereas the district-level respondent underscored field constraints such as human resource shortages, cold chain maintenance, and campaign fatigue. Together, these findings reveal a consistent recognition of digital potential, tempered by practical implementation gaps that require joint planning and targeted capacity investments.

#### 2.2.3 Co-Creation and Customisation of Tool Functionalities

The project adopted a co-creation approach, engaging EPI managers, vaccinators, and analysts from the start to ensure alignment with field realities. RCCA data were jointly analysed to refine tool features, enabling real-time syncing and enhanced visualisation through Power BI dashboards with contextual tooltips and decision-support visuals. On-site testing and continuous feedback in Lahore guided iterative refinement and improved user experience. This adaptive, user-centred design, built ownership and accountability, ensuring the tool’s relevance, usability, and sustainability within the immunisation ecosystem. By embedding stakeholder perspectives, the project strengthened both the digital platform and institutional readiness for scale-up.

### 2.3 Results – Fidelity, Acceptability, and Sustainability

#### 2.3.1 Fidelity Tracking and Stakeholder Feedback

##### 2.3.1.1 Orientation and Capacity Building

The fidelity tracking process was an integral component of the pilot implementation, ensuring that the intervention was delivered as intended while remaining responsive to field realities. Key activities included orientation and training of EPI and MIS teams at district and provincial levels on mobile app use, real-time field observation, stakeholder feedback loops, and adaptive modifications based on operational insights. Furthermore, front-line field workers nominated by District Lahore EPI team were provided comprehensive training on field methodology, use of customised mobile App to collect data from the selected union council of Lahore.

##### 2.3.1.2 Real-Time Field Observation and Stakeholder Feedback

Fidelity tracking involved on-site observation and hand-holding of field teams to assess real-time data syncing, functionality of mobile applications, and error-handling procedures. Stakeholder feedback guided enhancements in usability and functionality. The UC selection logic was optimised for local relevance, tooltips and visual guides were added to improve comprehension, and mobile data entry was streamlined to reduce errors. Integration of data visualisation with actionable planning allowed managers to identify coverage gaps and monitor corrective actions in real time.

##### 2.3.1.3 Adaptive Modifications

Adaptive technology enhancements included mobile-responsive dashboards, real-time data capture and review, automated 30-minute data refresh, improved mobile UI, and demonstrations to illustrate readiness for field deployment. Iterative refinement aligned the tool with operational realities, supporting timely action, accountability, and improved immunisation outcomes.

#### 2.3.2 Institutional Capacity and Sustainability

Punjab’s EPI Program has demonstrated a progressive and adaptive approach to digital health innovations in the past decade, most notably through the province-wide adoption of e-Vaccs and NEIR-EMR pilots. These experiences reveal several enabling factors and some systemic gaps that will influence sustained uptake of the newer digital monitoring and planning intervention.

Enablers include proven adoption capacity, established digital literacy, data-use culture, and stakeholder co-creation fostering buy-in. However, potential challenges that can influence the sustainability include competing priorities, administrative turnover, and dependence on external technical support, emphasising the need for internal capacity for tool maintenance and training.

## 3. Discussion

This study demonstrates the feasibility of embedding a digital monitoring and action planning tool within Pakistan’s immunisation system. The findings affirm the importance of co-creation in digital health interventions, as stakeholder engagement from inception to dissemination was crucial in ensuring operational relevance, ownership, and adoption. This aligns with broader evidence from literature, for instance, the study of Zindagi Mehfooz EIR in Sindh, found that along with system deployment, qualitative interviews revealed acceptance and utility when field actors felt engaged [16].

When compared to existing platforms such as e-Vaccs, NEIR/EMR, and EPI-MIS, the distinctive contribution of this tool lies in its integration of data visualisation with actionable planning. Whereas prior platforms were effective in monitoring performance and recording immunisation data, they lacked the capacity to generate and track targeted action plans at the UC level. For example, Sullivan’s case report on immunisation information systems in Sindh highlights that the Sindh immunisation system the current immunisation information system is electronic from the district to the provincial level but paper-based on the household to the district level [17]. This innovation thus fills a critical gap in enabling managers to not only identify coverage gaps but also to initiate and monitor corrective actions in real time.

The pilot provided valuable insights into the enablers and barriers of digital health adoption in resource-constrained settings. Key enablers included Punjab’s EPI program’s strong culture of accountability, prior exposure to digital tools, and stakeholder buy-in achieved through co-creation. These align with regional evidence showing that training, supportive supervision, and end-user engagement are critical for successful digital adoption [18]. Barriers included leadership turnover, inadequate training, limited digital literacy, and competing priorities, which mirror challenges documented in Bangladesh [19]. Scalability is ensured through capacity building of the EPI team and technology transfer, while addressing the remaining barriers will be essential to ensure sustainable scale-up.

Policy implications are significant. The study highlights the need for policy endorsement at provincial and national levels to institutionalise the tool within routine immunisation workflows. This is consistent with WHO and Gavi guidance, which emphasises integrating digital tools into routine workflows, providing long-term support, and ensuring interoperability for sustainability [20]. Sustained resource commitments, integration with existing digital platforms, and structured capacity-building initiatives will be essential. From a health systems perspective, the intervention demonstrates how digital innovations can advance equity by making zero-dose and under-immunised children more visible to program managers.

Limitations of the study include its narrow geographic scope, and reliance on qualitative feedback for assessing acceptability and usability. Future research should examine the cost-effectiveness of scaling the tool, its long-term impact on immunisation coverage, and the feasibility of utilising this data into action planning. Despite these limitations, the pilot provides compelling evidence of feasibility and policy relevance.

### 3.1 Pathway to Scale-up & Sustainability

“The Mainstream Initiative follows a stepwise pathway (Fig 11) starting from identifying immunisation gaps among zero-dose and under-immunised children, introducing a digital innovation (mobile app + dashboard), testing its implementation for accountability and responsiveness, and finally assessing the potential for scalability and sustainability. This progression reflects how the intervention moves from problem recognition to long-term system integration.”

**Fig 11:**
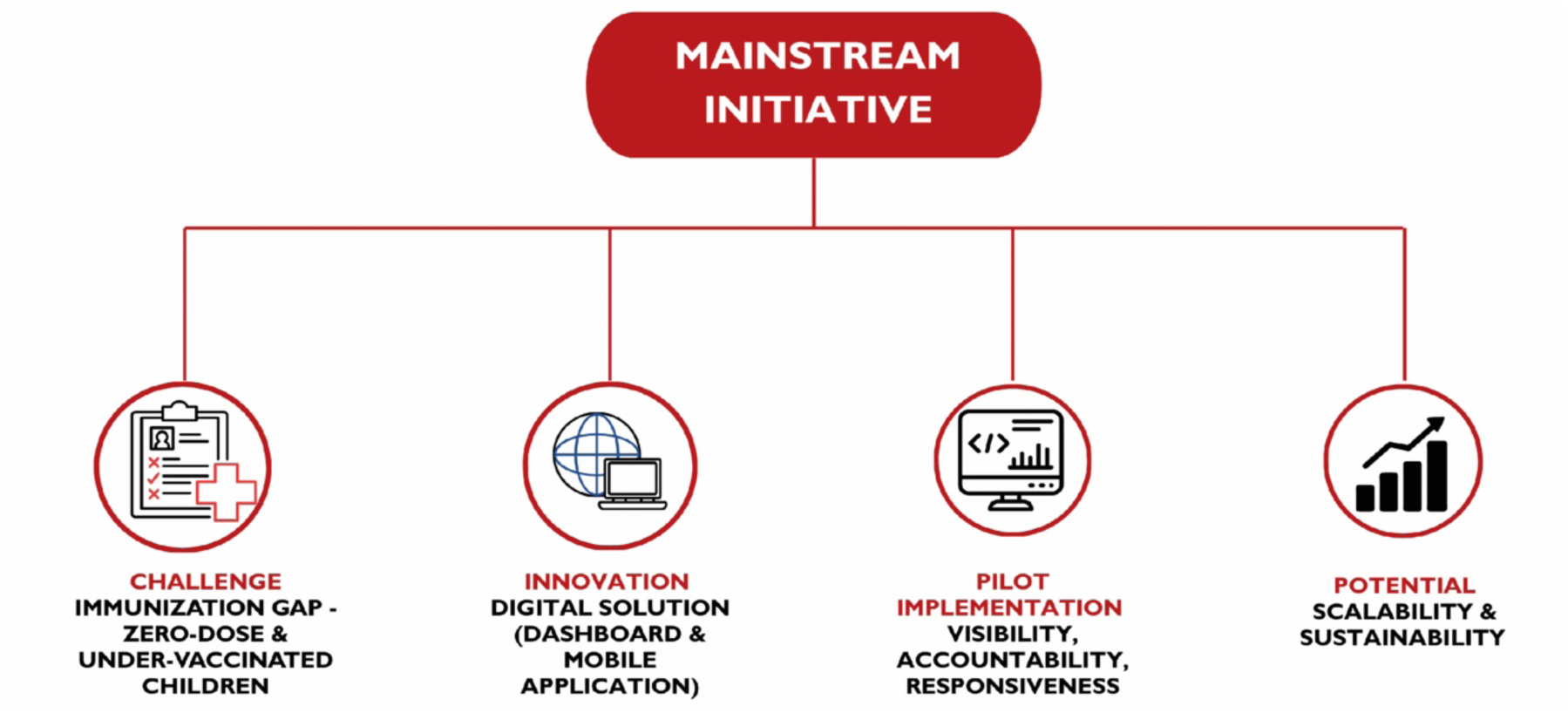
**Pathway to Scale-up and Sustainability**

The design visually illustrates the journey of the Mainstream Initiative in addressing immunisation gaps through digital innovation. It begins by highlighting the challenge of zero-dose and under-immunised children, represented as the core problem. This mirrors regional evidence: for example, in South Asia an estimated 3 million children remain un-or under-vaccinated, emphasising the equity challenge [21]. The pathway then transitions to the innovation; a digital solution combining a mobile application with a dashboard for frontline and supervisory use. This innovation leads into the implementation phase, emphasising accountability, accessibility, and responsiveness as key attributes tested through field deployment. Finally, the design points to the potential for scalability and sustainability, underscoring policy alignment and system integration as essential for long-term impact. The vertical flow reinforces the idea of a stepwise progression, from problem identification, to innovation, to testing, and ultimately to sustainable scaling, mirroring the objectives, findings, and lessons learned.

### 3.2 Conclusions and Recommendations

The Digital Monitoring and Action Planning Tool piloted under the MAINSTREAM Initiative demonstrated feasibility, acceptability, and potential for scalability. By integrating real-time data capture, geo-spatial mapping, and accountability mechanisms, the tool directly addressed persistent gaps in Pakistan’s immunisation program. The co-creation process ensured ownership, adaptability, and user-centred design, enhancing the likelihood of sustained adoption. With appropriate policy support, system integration, and capacity-building, the tool has the potential to transform immunisation monitoring and action planning across Punjab and beyond, contributing to improved equity and responsiveness in health systems. To support the institutionalisation and scalability of the digital health solution, the following recommendations are proposed:

#### 3.2.1 Establish Continuous Feedback Loops

Formal mechanisms should be developed to capture and share field-level observations, user experiences, and operational insights. These feedback loops will inform the iterative refinement of digital tools, research strategies, and implementation protocols.

#### 3.2.2 Institutionalise Knowledge and Skill Transfer

Dedicated efforts should be made to embed technical knowledge and digital competencies within the EPI system. This includes transferring the capacity to interpret digital data outputs, apply research findings, and operate the technology independently.

#### 3.2.3 Implement Structured Training and Refresher Programs

Comprehensive training modules and periodic refresher sessions should be designed and delivered to EPI managers and relevant staff (e.g. Vaccinators). These should focus on maximising the use of smartphone App, digital dashboards, interpreting analytics, and translating data insights into actionable plans and tracking outcome.

#### 3.2.4 Promote Transparent and Real-Time Communication

An enabling environment must be fostered for ongoing collaboration between researchers and implementers. Digital platforms should be leveraged to ensure timely, transparent, and bidirectional communication among stakeholders, enabling informed and coordinated decision-making.

Looking ahead, the digital monitoring and action planning tool, if embedded into routine immunisation systems with adequate capacity building, interoperability frameworks, and policy support, holds significant promise for achieving sustained improvements in vaccine coverage, particularly in under-reached and high-risk areas. It presents a scalable model that can be adapted across districts/provinces to advance Pakistan’s immunisation equity and system efficiency goals.

## 4. Methodology and Material

### 4.1 Study Design

An embedded implementation research (IR) design was employed, incorporating both primary and secondary data. This approach emphasised the integration of research within programmatic processes, ensuring that insights generated were directly applicable to decision-making and action planning. The design involved a combination of desk review, key informant interviews (KIIs), focus group discussions (FGDs), and pilot testing.

### 4.2 Geographical Scope and Study Sample

The study was conducted in the province of Punjab and was co-created with their Expanded Programme on Immunisation. The intervention was piloted in District Lahore, with data collected from 60 households across six clusters of a Union Council. Union Councils (UCs) with high concentrations of zero-dose children, low Penta-3 coverage, and urban slum populations were prioritised for the pilot.

### 4.3 Data Collection

Multiple sources of data informed the implementation process. Desk review mapped the immunisation landscape and digital health ecosystem in Punjab. Technical Team of Contech conducted KIIs and FGDs with provincial and district managers, supervisors, vaccinators, and community representatives to capture perspectives on challenges and needs. During the pilot periods i.e. from October 2024 to September 2025, data was collected by the vaccinators and visualised by the supervisors and managers. The pilot intervention generated evidence to assess fidelity, acceptability, adaptability and scalability potential.

### 4.4 Data Analysis

Data were analysed using a mixed-methods approach that combined quantitative and qualitative techniques to comprehensively assess the digital monitoring and action planning tool. Quantitative data collected during pilot intervention through the mobile application from 60 households across six clusters of a Union Council were processed using advanced descriptive and spatial analytics within Power BI. These analyses enabled visualisation of vaccination coverage at the UC level, identification of zero-dose and dropout patterns, and exploration of key drivers of non-vaccination and missed opportunities. The Power BI dashboard facilitated mapping of low-coverage areas and generation of actionable insights for program managers. Qualitative data from KIIs and FGDs were subjected to thematic analysis to examine the tool’s acceptability, adoption, and fidelity, as well as barriers and enablers to its implementation. Findings from both quantitative and qualitative strands were triangulated to inform context-specific recommendations, strengthen feedback loops, and guide planning for scalability and sustainability.

### 4.5 Ethical Considerations

Ethical Clearance exemption was obtained from the National Bioethics Committee for Research (NBCR-1216 Exemption) as the study did not involve direct patient interventions. Informed consent, confidentiality, and equity principles were observed throughout the process.

## Data Availability

The datasets generated and analyzed during the current study are not publicly available due to the inclusion of potentially identifiable household-level and programmatic data. De-identified datasets may be made available from the corresponding author upon reasonable request and with permission from the relevant provincial health authorities, in accordance with ethical and data governance approvals.

## Contributorship

**Conceptualization:** Naeem uddin Mian, Mariam Zahid Malik

**Software Development & Visualization:** Usama Naveed, Khayyam Bin Saeed, Muhammad Sufyan Chaudhary

**Formal Analysis & Writing – Original Draft Preparation:** Muhammad Wasim Mirza, Mariam Zahid Malik, Umer Farooq Rana, Adeel Alvi

**Writing – Review & Editing:** Maria Waheed, Amna Sattar

**Supervision:** Zahid Memon, Wardah Ahmed, Ammara Ali, Asma Ummad

## References

1. World Health Organization. Immunisation [Internet]. Geneva: World Health Organization; n.d. [cited 2025 Nov 11]. Available from: https://www.who.int/news-room/facts-in-pictures/detail/immunisation

2. Haq. Z, Shaikh BT, Tran N, Hafeez A, Ghaffar A. System within systems: challenges and opportunities for the Expanded Programme on Immunisation in Pakistan. Health research policy and systems. 2019 May 17;17(1):51.

3. Soofi SB, Hussain I, Kazi MA, Khan A, Umer M, Feroz K, Ahmed I, Rhoda DA, Bhutta ZA. Third-Party Verification Immunisation Coverage Survey (TPVICS) – 2023 [Internet]. Karachi: Aga Khan University; 2023 [cited 2025 Nov 11]. Available from: https://ecommons.aku.edu/pakistan_coe-wch_survey_report/2

4. Hussain I, Khan A, Rhoda DA, Ahmed I, Umer M, Ansari U, Shah MA, Yunus S, Brustrom J, Oelrichs R, Soofi SB, Bhutta ZA. Routine immunisation coverage and immunisation card retention in Pakistan: results from a cross-sectional national survey. Pediatr Infect Dis J. 2023 Mar;42(3):260–70. doi:10.1097/INF.0000000000003804

5. Punjab Information Technology Board (PITB). Punjab e-Vaccs program brings remarkable improvement. [Internet]. Lahore: PITB; 2015 Dec 5. Available from: https://www.pitb.gov.pk

6. Punjab Information Technology Board. PITB’s e-Vaccs system has improved vaccine coverage from 22% to 92% in three years [Internet]. Lahore: PITB; 2018 Jul 3 [cited 2025 Nov 11]. Available from: https://www.pitb.gov.pk

7. Zaidi S, Hussain SS, Riaz A, Rabbani F, Nisar MI, Khan AJ. Operability, acceptability, and usefulness of a mobile app to track routine immunisation performance in rural Pakistan: Interview study among vaccinators and key informants. JMIR Mhealth Uhealth. 2020;8(2):e16081. doi:10.2196/16081

8. Expanded Programme on Immunisation Punjab. NEIR integration with Punjab EMR vaccination module [Facebook post]. Lahore: EPI Punjab; 2024 [cited 2025 Nov 11]. Available from: https://www.facebook.com/EPIPakistanOfficial/posts/the-federaldirectorateofimmunisation-the-expanded-programme-on-immunisation-paki/866621208943819/

9. Punjab Health & Population Department. Expanded Program on Immunisation Punjab [Internet]. Lahore: Government of Punjab; n.d. [cited 2025 Nov 10]. Available from: https://neir.epimis.pk

10. Gavi, The Vaccine Alliance. How Punjab in Pakistan boosted its vaccination coverage. Vaccines Work. [Internet]. VaccinesWork; n.d. [cited 2025 Aug 16]. Available from: https://www.gavi.org/vaccineswork/how-punjab-pakistan-boosted-its-vaccination-coverage.

11. Federal EPI, Government of Pakistan. Standardization of Pakistan DLI 4: Vaccine Logistics Management Information System (vLMIS) [Press release]. [cited 2025 Oct 20]. Available from: https://lmis.gov.pk

12. United States Agency for International Development (USAID), Federal EPI Pakistan. Scaling up vLMIS across districts, including Punjab [Project document]. n.d. [cited 2025 Oct 22]. Available from: https://www.measureevaluation.org.

13. USAID DELIVER Project, Ministry of National Health Services. Improving Pakistan’s vaccine supply chain using vLMIS: Tracking consumption, cold chain, and stock data [User manual]. n.d. [cited 2025 Nov 11]. Available from: https://lmis.gov.pk

14. Federal EPI, Government of Pakistan; MEASURE Evaluation. Standardization of Pakistan DLI 4: Vaccine Logistics Management Information System (vLMIS) [Internet]. 2018. [cited 2025 Oct 22]. Available from: https://www.emro.who.int

15. Global Polio Eradication Initiative. Assessing Vaccination Coverage Levels Using Clustered Lot Quality Assurance Sampling: Field Manual, 2012. [cited 2025 June 16]. Available from: https://polioeradication.org/wp-content/uploads/2016/09/Assessing-Vaccination-Coverage-Levels-Using-Clustered-LQAS_Apr2012_EN.pdf. Accessed 11 February 2025.

16. Mechael P, Gilani S, Ahmad A, LeFevre A, Mohan D, Memon A, Shah MT, Siddiqi DA, Chandir S, Soundardjee R. Evaluating the “Zindagi Mehfooz” electronic immunisation registry and suite of digital health interventions to improve the coverage and timeliness of immunisation services in Sindh, Pakistan: mixed methods study. Journal of medical Internet research. 2024 Oct 11;26:e52792.

17. Sullivan E, Masood T, Javed W, Bagshaw K, Ollis S, Regmi P, Gardezi SM. Electronic immunisation information systems: a case report of lessons learned from implementation in Pakistan. Mhealth. 2020 Jul 5;6:31.

18. Kazi AM, Qazi SA, Ahsan N, Khawaja S, Sameen F, Saqib M, Khan Mughal MA, Wajidali Z, Ali S, Ahmed RM, Kalimuddin H. Current challenges of digital health interventions in Pakistan: mixed methods analysis. Journal of medical Internet research. 2020 Sep 3;22(9):e21691.

19. Taher A, Shimul MM, Khan S, Khandker S. Adoption challenges of digital transformation of human resource management in Bangladesh’s healthcare system: a cross-sectional mixed-methods evaluation. BMC Health Services Research. 2025 Oct 21;25(1):1383.

20. DHIS2. Improving National Immunisation Program Impact with DHIS2. [Internet]. Oslo: DHIS2; n.d. [cited 2025 November 11]. Available from: https://dhis2.org/immunisation/

21. UNICEF Regional Office for South Asia. Immunisation Regional Snapshot 2022: South Asia [Internet]. Kathmandu: UNICEF ROSA; 2022 [cited 2025 Nov 11]. Available from: https://www.unicef.org/rosa/reports/immunisation-regional-snapshot-2022-south-asia

